# Hypoglycemic Events in Diabetic Patients under Non-insulin Regimens During Ramadan: A Frequentist Network Meta-Analysis

**DOI:** 10.1101/2022.05.28.22275730

**Authors:** Salah Eddine O. Kacimi, Mahnoor Sukaina, Anas Elgenidy, Ramadan Abdelmoez Farahat, Huzaifa A. Cheema, Amira Y. Benmelouka, Dina M. Awad, Hadj Ahmed Belaouni, Moustafa K.E Abdelli, Anisse Tidjane, Nabil Smain Mesli, Jaffer Shah, Mounir Ould Setti, Ahmed M. Afifi, Sherief Ghozy, the Ramadan Diabetes Research Group (RDRG) Collaborators

## Abstract

**Background:** Diabetic Muslims who choose to fast during Ramadan encounter major risks such as hyperglycemia, hypoglycemia, diabetic ketoacidosis, and dehydration. Recently, newer antidiabetic agents have been found to be less likely to cause hypoglycemic emergencies. This meta-analysis aimed to present collective and conclusive results from major randomized controlled trials (RCTs) to determine the risk of hypoglycemia among patients taking oral antidiabetics during Ramadan.

**Methods:** We searched PubMed, Web of Science, and Google Scholar for RCTs. We performed a frequentist network meta-analysis using the “netmeta” package of R software version 4.1.1 to investigate the risk of developing hypoglycemia after taking oral antidiabetic drugs during Ramadan.

**Results:** Nine RCTs with a total of 3464 patients were included in the final analysis. In the comparison of all antidiabetic drug classes with sulfonylureas, SGLT-2 inhibitors were associated with the lowest hypoglycemic risk (RR, 0.18; 95% CI, 0.04-0.78; P-score, 0.909), followed by GLP-1 agonists (RR, 0.31; 95% CI, 0.17-0.56; P-score, 0.799), and DDP-4 inhibitors (RR, 0.57; 95% CI, 0.43-0.75; P-score, 0.483). When comparing individual drugs, dapagliflozin was associated with the lowest hypoglycemic risk (RR, 0.18; 95% CI, 0.04-0.78; P-score, 0.874), followed by lixisenatide (RR, 0.25; 95% CI, 0.09-0.71; P-score, 0.813), liraglutide (RR, 0.34; 95% CI, 0.17-0.69; P-score, 0.715), and sitagliptin (RR. 0.51; 95% CI, 0.37-0.71; P-score, 0.515).

**Conclusion:** SGLT-2 inhibitors are associated with the least documented hypoglycemic events and adverse outcomes compared with other oral hypoglycemic drugs. These findings could have considerable public health and clinical implications when extrapolated to the global Muslim population with a similar clinical background.

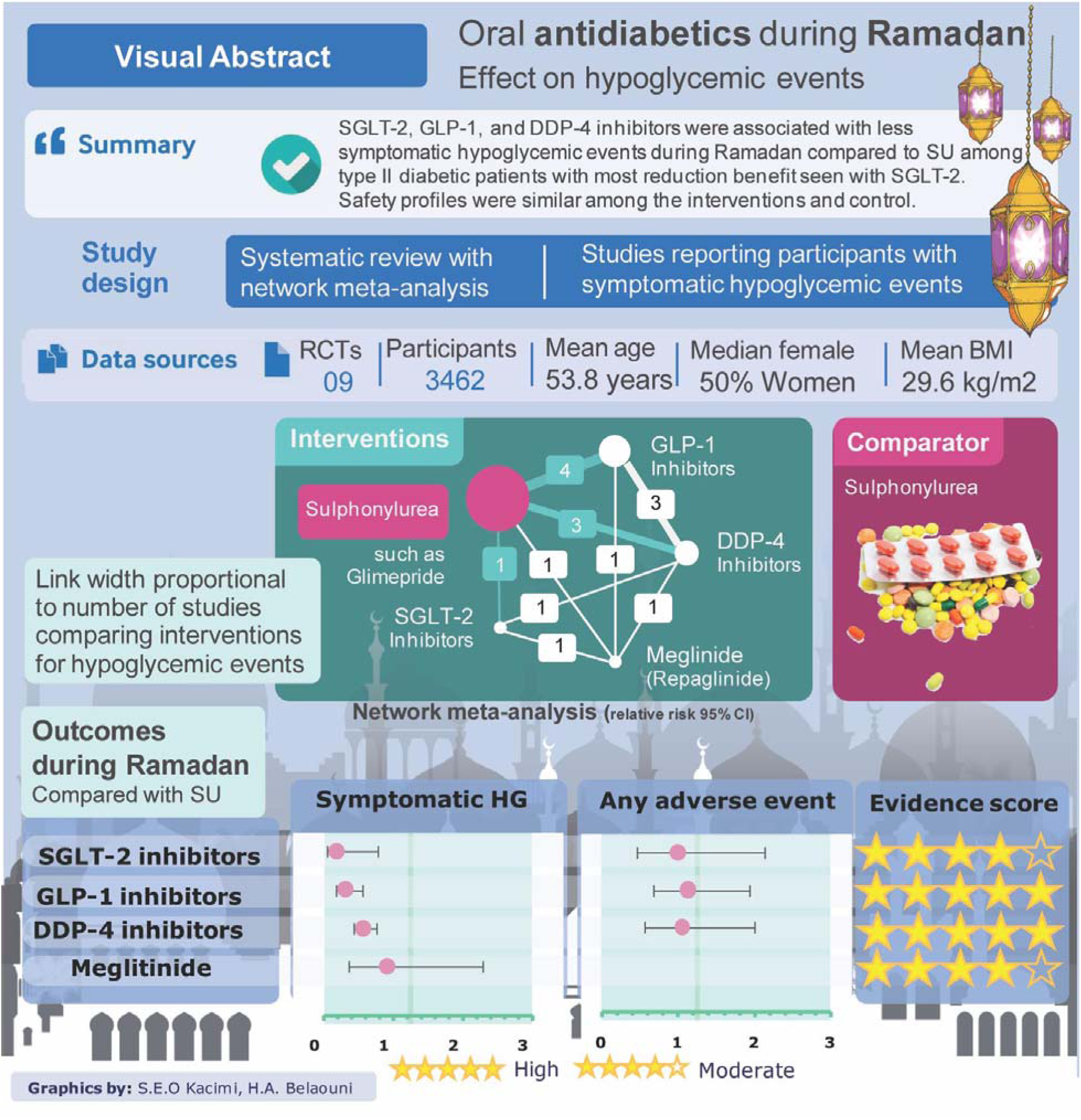

## Introduction

Fasting is one of the basic principles of Islam, a religion that over a billion people practice. Healthy Muslim adults must cease eating and drinking from sunrise to sunset during Ramadan, the 9^th^ lunar month in the Islamic calendar [1,2]. Presently, there are more than 150 million Muslim diabetics globally [3], and the number of diabetics worldwide is expected to surpass 360 million by 2030 [2]. Glycemic control in diabetic patients is essential. This is to avoid acute hyperosmolar non-ketotic coma, chronic diabetic complications such as diabetic retinopathy in microvascular changes, and other macrovascular complications [1].

However, maintaining glycemic control may be difficult, especially for diabetic Muslims who fast during the holy month of Ramadan.[4] Therein lies the major and serious complication of hypoglycemia associated with fasting and oral hypoglycemic antidiabetic such as meglitinides and sulfonylureas [1,5,6]. The Epidemiology of Diabetes and Ramadan (EPIDIAR) demonstrated that around 78.5% of Muslim diabetic patients who fast did not refrain from fasting for at least 15 days. Furthermore, an alteration in physical activity and changes in insulin dosage and other oral hypoglycemic drugs were observed to increase severe hypoglycemic events [7].

In recent times, newer antidiabetic agents have been introduced which are less likely to cause hypoglycemic emergencies. Such antidiabetics include dipeptidyl peptidase-4 (DPP-4) inhibitors and sodium-glucose co-transporter-2(SGLT-2) inhibitors. Crucially, the role of SGLT-2 inhibitors has been rarely elaborated in a network meta-analysis to show efficacy in fasting diabetic patients. Currently, most studies have shown that hypoglycemia during Ramadan can be curbed by shifting the therapy from insulin or other oral hypoglycemics to DPP-4 inhibitors or metformin [1,5,6]. A network meta-analysis by Lee et al. showed that newer antidiabetics exhibit appropriate glycemic control and lower hypoglycemic complications than sulfonylureas [8]. Moreover, it also highlighted that incretin-based mimetics manifest 1.5 times lesser hypoglycemic events [8]. Likewise, a recent meta-analysis demonstrated that DPP-4 and glucagon-like peptide (GLP-1) agonist therapy significantly improves HbA1c levels amongst fasting diabetic patients during Ramadan [9]. Other studies have also shown that Ramadan-focused diabetes education helps create awareness among the population, ensuring a decline in HbA1c levels in fasting patients [10].

This systematic review and meta-analysis aimed to present collective and conclusive results of major randomized controlled trials (RCTs) regarding hypoglycemic events associated with antidiabetic medication in the fasting population during Ramadan. Diabetic emergencies, if not treated timely, can lead to fatal outcomes. Lack of awareness and standard therapy hinders seeking and continuing treatment. The rationale of our study is to fill the literature gap and provide the data that helps create a standardized therapy for fasting people with diabetes during Ramadan.

## Methods

This meta-analysis was conducted according to the Preferred Reporting Items for Systematic Review and Meta-analyses (PRISMA) statement recommendations [11]. A systematic search was done to select all the trials that correspond to the following criteria: 1) population: diabetics fasting during Ramadan; 2) intervention and comparator: any non-insulin oral antidiabetic agent compared with another oral agent; 3) outcome: hypoglycemic events; and 4) study design: RCTs.

### Literature search

The search was conducted on March 2022 in the following databases: MEDLINE (via PubMed), Scopus, Web of Science, and Science Direct using a search string relating to the following keywords: Ramadan, diabetes, oral antidiabetic agents, and hypoglycemia.

There were no restrictions on language, country, gender, race, or sample size among the published articles. Animal reports, reviews, letters, commentaries, conference abstracts/posters, case reports, case series, and trials including diabetic patients not fasting in Ramadan were excluded. Further, a manual search of reference lists of the included studies was done to retrieve any relevant studies.

### Screening and study selection

After removing duplicates, the title and abstract screening of the selected articles was performed by two independent reviewers. This step was followed by a full-text screening of the selected articles to verify the included population and the availability of the outcomes. When full texts were not available, we contacted the authors. Disagreements were resolved by a third senior researcher.

### Data extraction

Two independent authors performed the data extraction using a pre-performed excel sheet. Population demographics and data on hypoglycemic events during Ramadan were extracted. Disagreements were resolved through discussion with a senior researcher to find a consensus.

### Risk of bias

Two independent investigators evaluated the risk of bias in the included studies using the revised Cochrane Risk of Bias Tool for randomized controlled trials (RoB 2.0) [12]. RoB 2.0 addresses five domains: (1) bias arising from the randomization process; (2) bias due to deviations from intended interventions; (3) bias due to missing outcome data; (4) bias in measurement of the outcome; and (5) bias in the selection of the reported result. Each item was described as having a low, high, or unclear risk of bias. Each study’s overall risk of bias was described as low, moderate, or high risk, based on our judgments for all the items. A senior author resolved any discrepancies among reviewers by consensus.

### Statistical analysis

We conducted the analysis using the ‘netmeta’ package ver. 2.1-0 (https://cran.r-project.org/web/packages/netmeta/index.html) of R software version 4.1.1. We performed a frequentist network meta-analysis to investigate the risk of developing hypoglycemia after taking oral antidiabetic drugs during Ramadan. Fixed- or random-effects models were used to perform the network meta-analysis based on the level of heterogeneity/inconsistency [13]. When ≥10 studies were available, we built comparison-adjusted funnel plots to examine small study effects [13]. Funnel plot asymmetry was assessed with three tests (Begg–Mazumdar test, Egger’s regression, and Thompson–Sharp test). P values <0.1 were considered significant [14–16].

## Results

### Description of Eligible Trials

After screening, nine studies were selected for final analysis [17–25] **(Fig. 1)**. Five studies were multi-centric and conducted in several countries while the rest were conducted in the UK, Lebanon, Malaysia, India, and Malaysia. All included studies were RCTs with double arm comparisons with sulfonylureas **(Fig. 2)**. The total number of patients was 3464, with 1743 in the sulfonylurea treated group (mean age=53.6), and 1721 (mean age=53.9) in the intervention group. The intervention group included patients treated with the following drugs: sitagliptin, liraglutide, vildagliptin, lixisenatide and basal insulin, repaglinide, and dapagliflozin plus metformin. The mean BMIs for the intervention and sulfonylurea treated groups were 29.7 and 29.4, respectively (**Tab. 1**).

**Figure 1:**
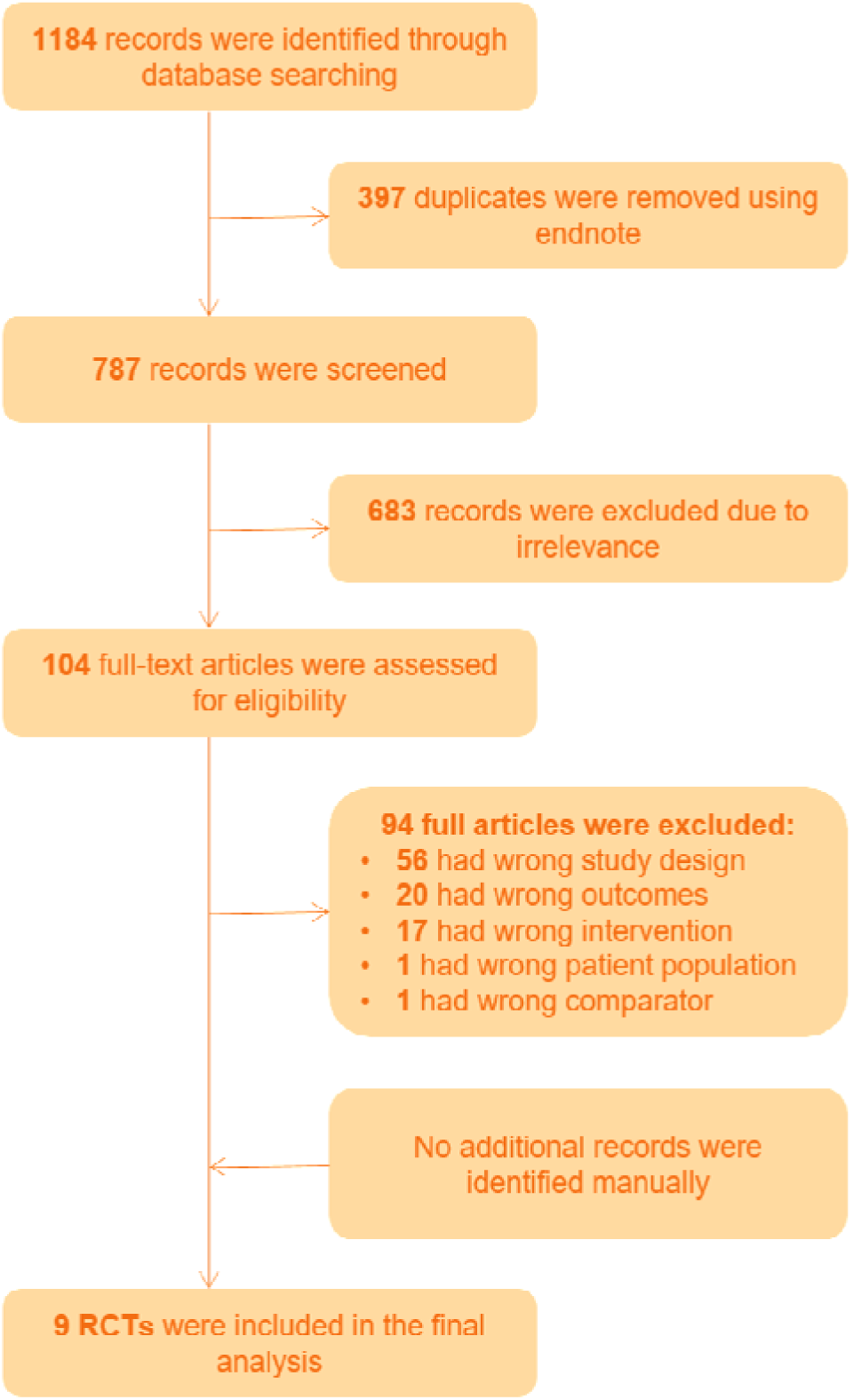
The PRISMA flow diagram of the study selection process.

**Figure 2:**
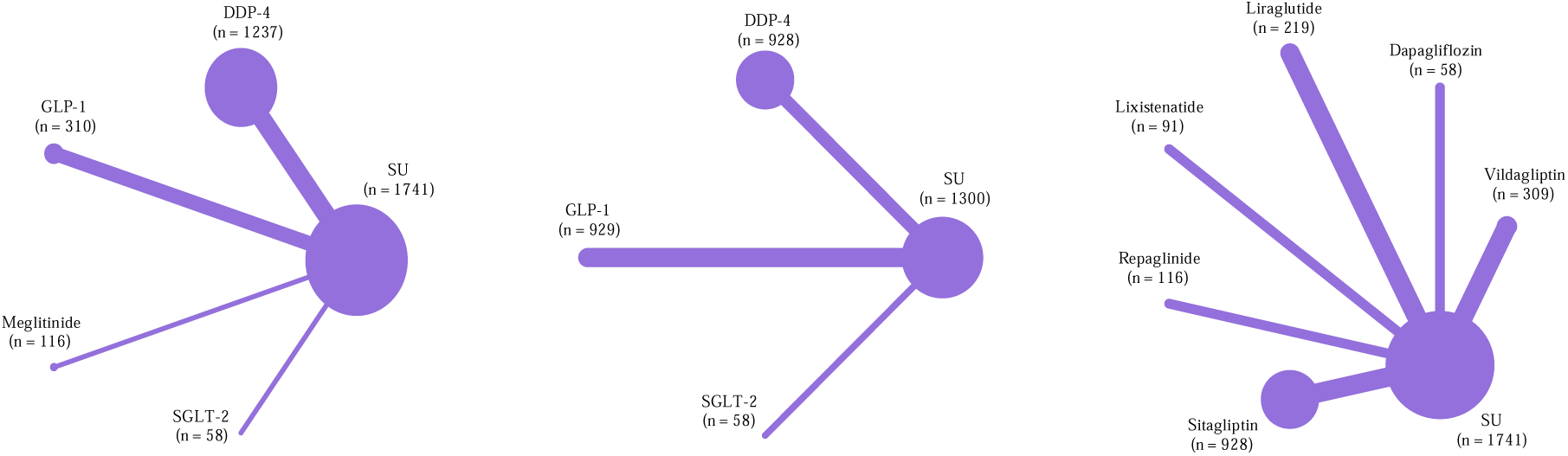
Network plots of all included studies for: i) antidiabetic drug class and symptomatic hypoglycemia, ii) antidiabetic drug class and any adverse effects, iii) drug name and symptomatic hypoglycemia.

**Table 1:** Basic characteristics of included articles

| Study ID | Intervention name | Intervention |  |  |  |  | Sulphonylurea |  |  |  |  |
| --- | --- | --- | --- | --- | --- | --- | --- | --- | --- | --- | --- |
|  |  | Total, n | Sympt. HG* | Age, mean (SD) | Females (%) | BMI, mean (SD) | Total, n | Sympt. HG | Age, mean (SD) | Females (%) | BMI, mean (SD) |
| Al Sifri et al. 2011 | Sitagliptin | 507 | 34 | 55 ± 11 | 238 (46.9) | 30.5 ± 5.7 | 514 | 68 | 55 ± 10 | 259 (50.39) | 30.5 ± 5.6 |
| Aravind et al. 2012 | Sitagliptin | 421 | 16 | 51.4 ± 9.9 | 213 (50.59) | 27.4 ± 6.0 | 427 | 31 | 50.7 ± 10.0 | 233 (54.57) | 27.5 ± 4.7 |
| Azar et al. 2016 | Liraglutide | 171 | 5 | 54.9 ± 9.27 | 86 (50.3) | 30.2 ± 5.37 | 170 | 16 | 54.0 ± 9.33 | 87 (51.2) | 31.4 ± 5.88 |
| Brady et al. 2013 | Liraglutide plus metformin | 47 | 12 | 51.5 ± 11.1 | 23 (48.94) | 33.0 ± 7.3 | 52 | 24 | 52.2 ± 10.7 | 26 (50) | 30.1 ± 4.3 |
| Hassanein et al. 2014 | Vildagliptin | 279 | 8 | 54.6 ± 9.3 | 147 (52.7) | 30.7 ± 5.0 | 278 | 19 | 54.3 ± 9.1 | 150 (54.0) | 31.1 ± 5.2 |
| Hassanein et al. 2019 | Lixisenatide +basal insulin | 92 | 3 | 52.6 ± 9.5 | 52 (56.5) | 29.7 ± 5.3 | 92 | 8 | 54.1 ± 10.6 | 49 (53.3) | 29.0 ± 4.3 |
| Mafauzy et al. 2002 | Repaglinide | 116 | 8 | 52.7 ± 7.4 | 29 (25) | 26.5 ± 2.5 | 119 | 9 | 54.5 ± 6.9 | 37 (31.1) | 26.8 ± 3.2 |
| Malha et al. 2014 | Vildagliptin | 30 | 19 | 57.0 ± 9.6 | NA | 29.49 ± 4.66 | 39 | 26 | 54.6 ± 9.2 | NA | 28.90 ± 4.49 |
| Wan Seman et al. 2016 | Dapagliflozin +metformin | 58 | 4 | 53 ± 9.1 | 23 (39.7) | 29.9 ± 4.84 | 52 | 15 | 56 ± 9.1 | 21 (40.4) | 29.64 ± 4.44 |

\*: Symptomatic

### Results of the Outcomes in NMA Hypoglycemia risk

Among the diabetic patients, the network of treatment comparisons for hypoglycemia was reported in nine studies [17–25]. The sulfonylurea was the well-connected group directly linked to all other treatments. In the comparison of all antidiabetic drug classes with sulfonylureas, SGLT-2 inhibitors were associated with the lowest hypoglycemic risk (RR, 0.18; 95% CI, 0.04-0.78; P-score, 0.909), followed by GLP-1 agonists (RR, 0.31; 95% CI, 0.17-0.56; P-score, 0.799), and DDP-4 inhibitors (RR, 0.57; 95% CI, 0.43-0.75; P-score, 0.483). Compared with each other, SGLT-2 inhibitors, GLP-1 agonists, DDP-4 inhibitors, and meglitinide did not reveal a statistically significant difference **(Fig. 3)**.

**Figure 3:**
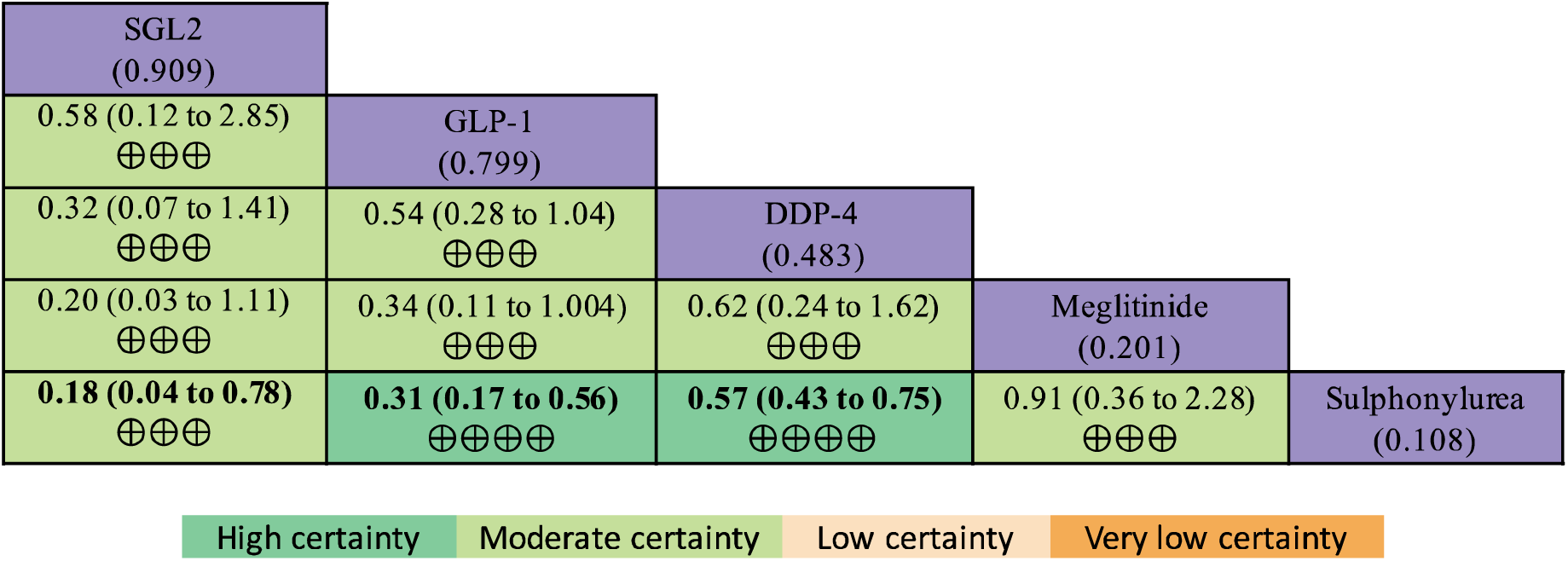
Non-insulin antidiabetics network meta-analysis results with corresponding GRADE (grading of recommendations, assessment, development, and evaluation) certainty of the evidence for symptomatic hypoglycemia. Values correspond to the relative risk of having at least one symptomatic hypoglycemic event when comparing columns and rows during Ramadan. Values in bold indicate a statistically significant treatment effect. Values under the treatment’s names correspond to the P-scores for the network ranking.

Dapagliflozin was associated with the lowest hypoglycemic risk (RR, 0.18; 95% CI, 0.04-0.78; P-score, 0.874), followed by lixisenatide (RR, 0.25; 95% CI, 0.09-0.71; P-score, 0.813), liraglutide (RR, 0.34; 95% CI, 0.17-0.69; P-score, 0.715), and sitagliptin (RR. 0.51; 95% CI, 0.37-0.71; P-score, 0.515), Ranking of the risk of hypoglycemia using p-score revealed dapagliflozin as the best, and sulfonylureas as the worst among treatments (P-score, 0.093). Compared with each other, dapagliflozin, lixisenatide, liraglutide, sitagliptin, vildagliptin, and repaglinide did not reveal a statistically significant difference **(Fig. 4)**.

**Figure 4:**
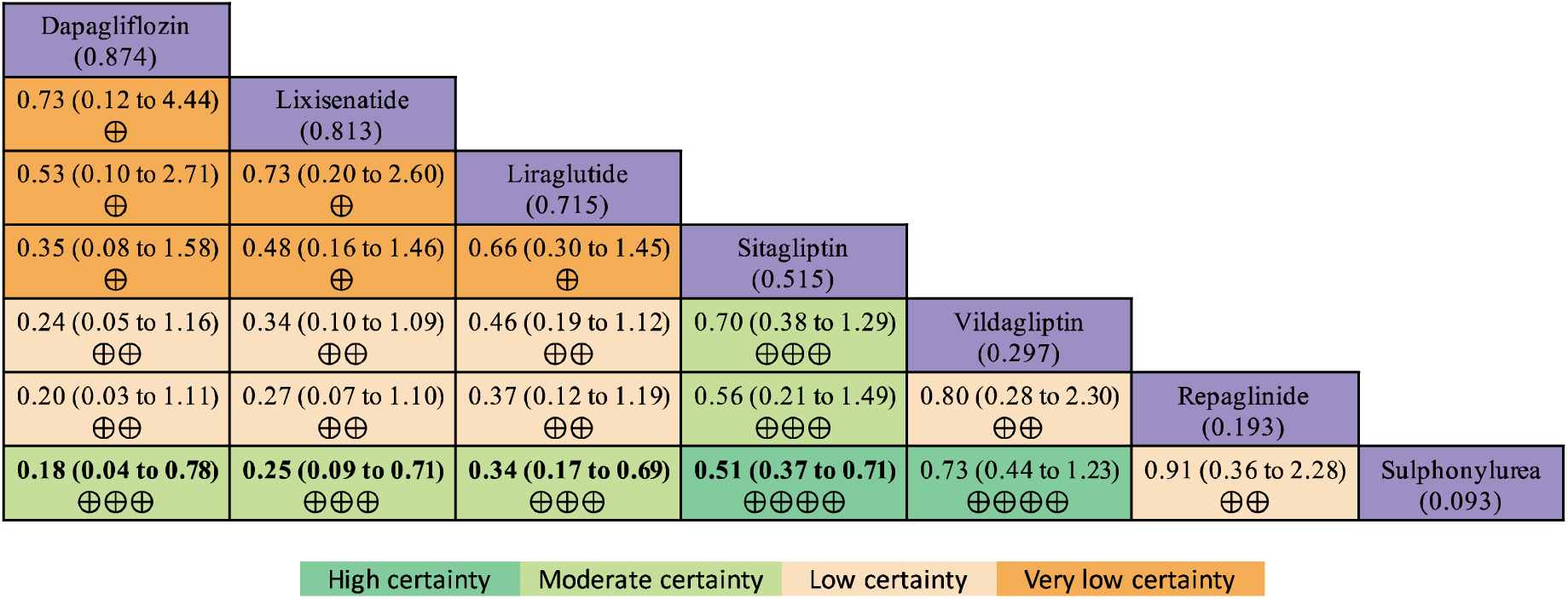
Non-insulin antidiabetics network meta-analysis results with corresponding GRADE (grading of recommendations, assessment, development, and evaluation) certainty of the evidence for symptomatic hypoglycemia. Values correspond to the relative risk of having at least one symptomatic hypoglycemic event when comparing columns and rows during Ramadan. Values in bold indicate a statistically significant treatment effect. Values under the treatment’s names correspond to the P-scores for the network ranking.

### Adverse effects

Comparing all treatments with sulfonylureas, there were almost no statistically significant differences regarding the relative risk of having any adverse event **(Fig. 5)**. However, ranking the risk of any adverse events using P-scores revealed sulfonylurea as the best (P-score, 0.59), with fewer adverse events, and GLP-1 agonists as the worst among treatments (P-score, 0.38). There was no statistically significant difference between the drug classes compared to each other.

**Figure 5:**
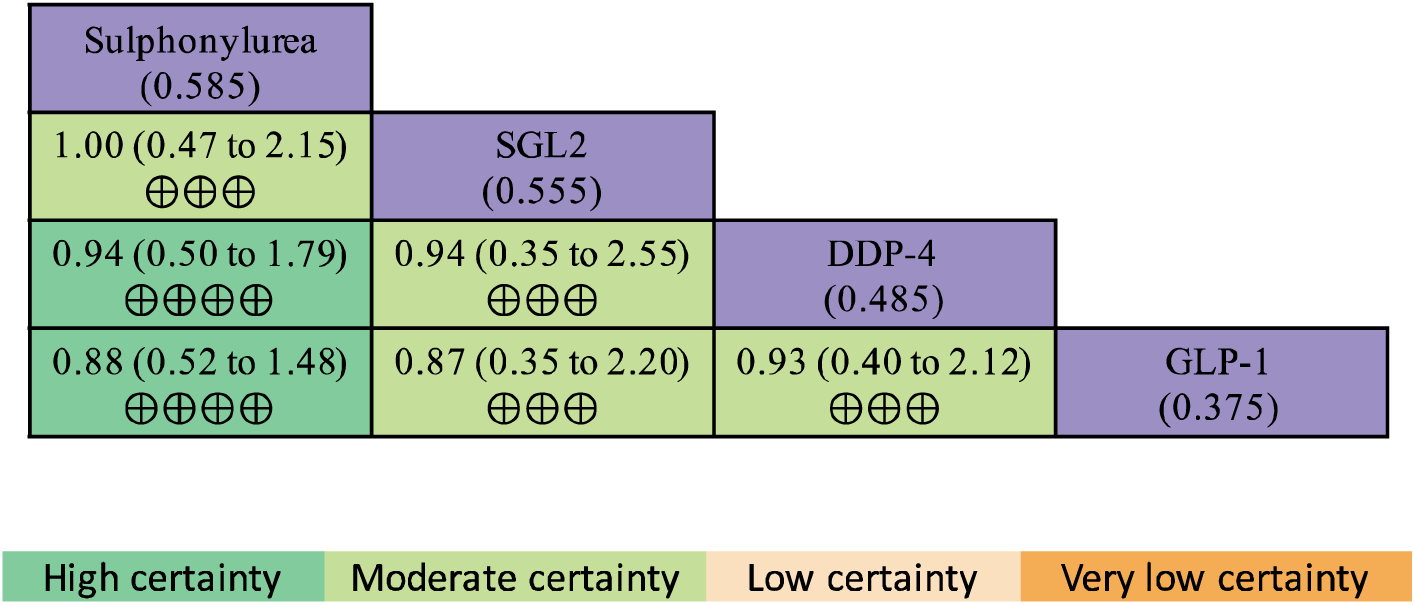
Non-insulin antidiabetics network meta-analysis results with corresponding GRADE (grading of recommendations, assessment, development, and evaluation) certainty of the evidence for any adverse events. Values correspond to the relative risk of having any adverse event when comparing column and row during Ramadan. Values in bold indicate a statistically significant treatment effect. Values under the treatment’s names correspond to the P-scores for the network ranking.

### Risk of Bias of Included Studies

The quality of included studies ranged from moderate to high quality according to the Cochrane Collaboration’s tool for assessing the risk of bias 2 (RoB 2). Regarding the randomization process, four studies were of low risk of bias [17,19,21,22], while five studies [18,20,23–25] were judged to have some concerns due to the lack of blinding patients and the study personnel.

All included trials had a low risk of bias in terms of deviations from the intended interventions, the missing outcome data, the measurement outcome bias, and the selection of the reported results bias. The detailed risk of bias domains by study ID, are reported in the supplement.

### Sensitivity Analyses

We compared the individual drugs of the sulfonylurea class with other antidiabetic drugs. The network of treatment comparisons for hypoglycemia consisted of eight individual nodes. Gliclazide was the well-connected group and directly linked to all other treatments.

In the comparison of all individual drugs with glibenclamide, liraglutide was associated with the lowest hypoglycemic risk (RR, 0.12; 95% CI, 0.03-0.49; P-score, 0.92), followed by lixisenatide (RR, 0.15; 95% CI, 0.04-0.62; P-score, 0.88), sitagliptin (RR, 0.43; 95% CI, 0.21-0.87; P-score, 0.60), vildagliptin (RR. 0.43; 95% CI, 0.12-1.48; P-score,0.57), gliclazide (RR, 0.61; 95% CI, 0.29-1.25; P-score, 0.40), glimepiride (RR. 0.74; 95% CI, 0.39-1.42; P-score,0.28), and repaglinide (RR, 0.95; 95% CI, 0.27-3.10; P-score, 0.23; **Fig. 6**). Ranking of the risk of any adverse events using p-score revealed liraglutide as the best, being with fewer hypoglycemic events, and glibenclamide as the worst among treatments (P-score, 0.117).

**Figure 6:**
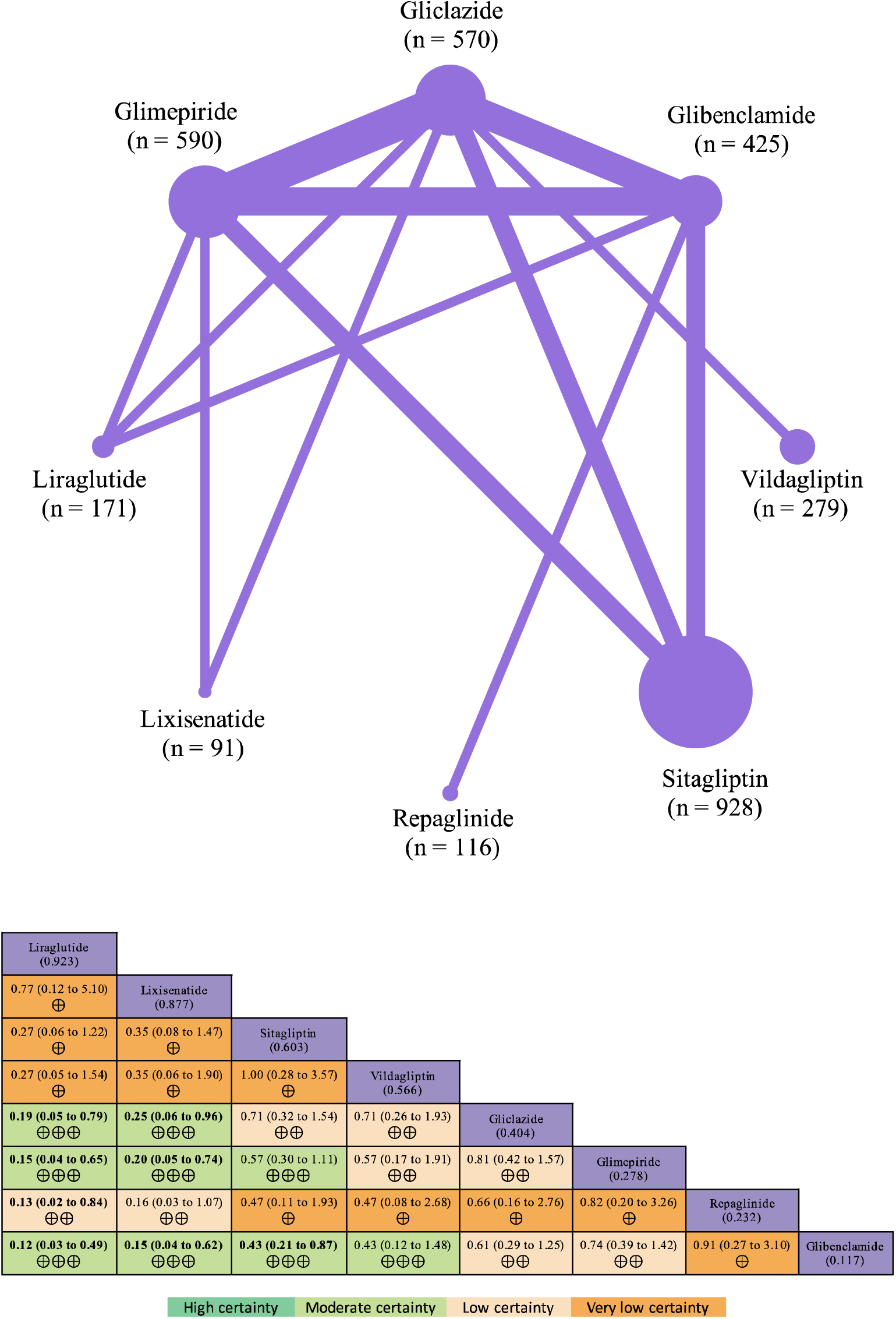
Network plots and sensitivity network meta-analysis results with corresponding GRADE (grading of recommendations, assessment, development, and evaluation) certainty of the evidence for symptomatic hypoglycemia. Values correspond to the relative risk of having at least one symptomatic hypoglycemic event when comparing columns and rows during Ramadan. Values in bold indicate a statistically significant treatment effect. Values under the treatment’s names correspond to the P-scores for the network ranking.

#### Among incretins mimetics

Liraglutide was better than lixisenatide, but there was no statistically significant difference regarding hypoglycemic events. Liraglutide and lixisenatide were better than **sulfonylureas** (gliclazide and glimepiride, glibenclamide), with statistically significant differences. Both Liraglutide and lixisenatide were better than **repaglinide**, but Liraglutide showed a statistically significant difference compared with repaglinide but not lixisenatide. Finally, Liraglutide and lixisenatide were better than **DPP-4 inhibitors** (sitagliptin, vildagliptin) with no statistically significant difference.

#### Among DPP-4 inhibitors

Sitagliptin was better than vildagliptin, with no statistically significant difference between them regarding the hypoglycemic events. Sitagliptin and vildagliptin were better than **sulfonylureas drugs** (Gliclazide and glimepiride, glibenclamide), with a statistically significant difference only between Sitagliptin and glibenclamide. Sitagliptin and vildagliptin were better than **repaglinide** with no statistically significant difference between them.

#### Among sulfonylureas

Gliclazide was better than glimepiride, and glibenclamide with no statistically significant difference between them regarding the hypoglycemic events. Both Gliclazide and glimepiride were better than **repaglinide**, on the other hand, repaglinide is better than glibenclamide. However, there was no statistically significant difference between them regarding the hypoglycemic events **(Fig. 6)**.

## Discussion

To the best of our knowledge, this is the first network meta-analysis of all newer oral hypoglycemic agents explicating hypoglycemic events in fasting diabetic patients during Ramadan. We found that SGLT-2 inhibitors are associated with the least documented hypoglycemic events and adverse outcomes followed by GLP-1 agonists, DPP-4 inhibitors, meglitinide and sulfonylureas. A recent meta-analysis by Gad et al. advocates the treatment efficacy of SGLT-2 inhibitors in Ramadan with fewer major adverse events [26]. Other meta-analyses by Shiju et al. and Loh et al.promote treatment with vildagliptin in high-risk patients, especially the elderly [27,28].

Original studies reporting that vildagliptin is associated with lesser hypoglycemic events during Ramadan coincide with our results [21,29,30]. However, when comparing both classes, vildagliptin has slighter lower efficacy as compared to sitagliptin, but non-significant better results than sulfonylureas. Gad et al. elucidated similar outcomes that favor the efficacy of SGLT-2 inhibitors, GLP-1 agonists, and DPP-4 inhibitors; however, the study does not ascertain the efficiency of newer oral hypoglycemic in the hierarchy of least hypoglycemic events.[9]Furthermore, the meta-analysis by Gad et al. approves both lixisenatide and liraglutide for fasting diabetic patients in Ramadan like our study. Similarly, a meta-analysis by Gray et al. presented the clinical efficacy of liraglutide [31]. We demonstrate that lixisenatide shows better efficacy than liraglutide; however, both lower risk of symptomatic hypoglycemia compared with sulfonylureas.

We acknowledge the statement by Gray et al. that the majority of trials and observational studies favor the treatment efficacy of DPP-4 inhibitors over sulfonylureas in fasting T2D patients [31]. Our meta-analysis shows the possible efficacy of SGLT-2 inhibitors over GLP-1 agonists and the aforementioned drugs over DPP-4 inhibitors, although all these drug classes are advantageous in preventing hypoglycemic events in fasting diabetics. However, like Gray et el. we recommend that in the future, more studies should evaluate and compare SGLT-2 inhibitors, GLP-1 agonists, and DPP-4 inhibitors in terms of efficacy, effectiveness, primary and secondary outcomes, and safety outcomes. A study by Lee et al. showed that DPP-4 inhibitors can reduce the occurrence of hypoglycemic events in people who fast during Ramadan when compared to sulfonylureas [8]. However, it was inconclusive regarding meglitinides due to the lack of relevant data on people observing fasting. The study by Mbanya et al. compared sulfonylureas and found gliclazide to be the better choice of drug as compared with glimepiride which is better when compared to metformin [32]. On the contrary, the observation study by Bonakdaran and Khajeh-Dalouie concluded that sulfonylureas have a significantly higher incidence of hypoglycemic events as compared to metformin [33]. The LIRA-Ramadan study by Azar et al. concluded that using liraglutide can have significantly better outcomes and lesser complications, including hypoglycemic events, compared with sulfonylureas [23]. Similarly, our study shows that GLP-1 agonists have better outcomes when compared with DPP-4 inhibitors, meglitinides, and sulfonylureas. John et al. discussed that SGLT-2 inhibitors provide lesser risk or severity of hypoglycemia in patients living in warm climate conditions [34]. The majority of Muslims live in warm climatic conditions, and therefore fasting in Ramadan has better results when people are using SGLT-2 inhibitors and may reduce the risk of hypoglycemia.

Currently, amidst the COVID-19 pandemic, Tootee et al. discussed that SGLT-2 inhibitors be avoided due to the heightened risk of diabetic ketoacidosis (DKA) and dehydration as COVID-19 infection may cause compromised immunity [35]. Hassanein et al., in their CRATOS study, concluded that SGLT-2 inhibitors are a safer option for people fasting in Ramadan as they have a lower incidence of hypoglycemic events [36]. However, they still have the risk of volume depletion due to osmotic diuresis, especially in the elderly group of patients [37].

We recommend using newer technology, non-invasive devices over the regular finger prick glucometers to monitor glucose levels throughout the day in fasting diabetics, which will aid in prick-free testing without voiding the fast and help recognize the best possible management regimen of diabetic drugs for individuals with different co-morbidities [38]. These devices can read glucose levels 24 hours a day and keep-up the track for up to two weeks.

### Strengths and Limitations

We presented a network meta-analysis of associations of different newer oral hypoglycemic drugs with the extent of hypoglycemia and adverse events from multicenter studies in different races and populations. Several limitations we observed include only hypoglycemic events as our outcome and not changes in HbA1c, weight loss, and changes in systolic and diastolic blood pressures. Moreover, we presented general adverse events and did not specify the most to least common adverse events associated with each drug class. We specified different drugs in each class, but we were only able to evaluate the efficacy of dapagliflozin in the class of SGLT-2 inhibitors. Another limitation that needs to be pointed out is the scarcity of studies on this topic with only nine trials matching our selection criteria.

## Data Availability

Data of this study are available upon reasonable request from the corresponding author.

## OTHER INFORMATION

### Availability of data, code, and other materials

Data of this study are available upon reasonable request from the corresponding author.

